# ESTIMATING THE HEALTH AND ECONOMIC EFFECTS OF THE VOLUNTARY SODIUM REDUCTION TARGETS IN BRAZIL: MICROSIMULATION ANALYSIS

**DOI:** 10.1101/2021.01.31.21250647

**Authors:** Eduardo Augusto Fernandes Nilson, Jonathan Pearson-Stuttard, Brendan Collins, Maria Guzman-Castillo, Simon Capewell, Martin O’Flaherty, Patrícia Constante Jaime, Chris Kypridemos

**Affiliations:** Department of Public Health, Policy & Systems, University of Liverpool, Liverpool, UK

**Keywords:** Sodium, sodium reduction, sodium targets, health economics, cardiovascular disease, hypertension, food policy, public health, global health

## Abstract

**Objective:** To analyse the potential health and economic impact of the voluntary sodium reduction targets in Brazil, from 2013 to 2032.

**Design:** Modelling study. A microsimulation approach of a close-to-reality synthetic population (IMPACT _*NCD BR*_) was used to evaluate the potential health benefits of setting voluntary upper limits for sodium content as part of Brazilian government strategy. The model estimates cardiovascular disease (CVD) deaths and cases prevented or postponed, and disease treatment costs.

Model inputs were informed by the 2013 National Health Survey, the 2008-2009 Household Budget Survey, and high-quality meta-analyses to inform model inputs. Costs included costs of the National Health System on CVD treatment and informal care costs.

**Setting:** Synthetic population with similar characteristics to the community dwelling population of Brazil.

**Participants:** Synthetic people with traits informed by the national surveys of Brazil.

**Main outcome measures:** Cardiovascular disease cases and deaths prevented or postponed by 2032, over a 20-year period (2013-2032), stratified by age and sex.

**Results:** Applying the voluntary sodium targets between 2013 and 2032 could prevent or postpone approximately 112,000 CVD cases (95% Uncertainty Intervals UI: 28,000 to 258,000) among men and 70,000 cases among women (95% UI: 16,000 to 167,000), and also prevent or postpone approximately 2,600 CVD deaths (95% UI: −1,000 to 11,000), 55% in men. The policy could also produce a net cost saving of approximately US$ 222 million (95% UI: US$ 53.6-524.4 million) in medical costs to the Brazilian National Health System for the treatment of CHD and stroke, and save approximately US$ 71 million (95% UI: US$ 17.1-166.9 million) in informal costs.

**Conclusions:** Brazilian voluntary sodium targets could generate substantial health and economic impacts. Further progress in lower, more comprehensive thresholds for sodium in foods and strategies for reducing other sodium sources could maximise the health and economic benefits to the population. This is the first IMPACT NCD microsimulation model adapted to a Latin American country and represents a big step forward for using models to inform policy in the region. The results indicate that sodium reduction targets must go further and faster in order to achieve national and international commitments.

**WHAT IS ALREADY KNOWN ON THIS TOPIC:**

- Public-private partnerships (PPPs), including voluntary targets for the reduction of critical nutrients, such as sodium, sugars and fats, through food reformulation, have been promoted as effective strategies for addressing dietary factors for non-communicable disease prevention.
- Salt (sodium chloride) intake is a leading dietary risk factor for cardiovascular disease (CVD) globally. Over 27 thousand deaths from coronary heart disease and stroke are attributable to excessive sodium intake in Brazil every year. About 20% of sodium in the Brazilian diet comes from industrialized foods, and over 70% come from added table salt and salt-based condiments.
- Since 2011, Brazil has implemented a voluntary approach for reducing sodium in processed and ultra-processed foods, including salt-based condiments. Nevertheless, national targets have not matched the targets of other countries in the Region of the Americas and globally, and target compliance has not been achieved across the entire Brazilian food market.

**WHAT THIS STUDY ADDS:**

- We estimated the impact of the current sodium reduction targets in Brazil, by analysing individual-level food category consumption and sodium intake.
- Using the first IMPACT NCD microsimulation model adapted to the Latin American context, we estimated that if applied between 2013 and 2032, the voluntary targets could potentially have prevented approximately 180,000 CVD cases and 2,500 CVD deaths. The case reductions might save approximately US$ 220 million in CVD-related medical costs (hospitalizations, outpatient and primary health care and pharmaceutical treatment) and some US$ 70 million in informal costs.
- More impactful sodium reductions in Brazil may not be achieved without more stringent and comprehensive targets; for instance, mandatory rather than voluntary policy formulation, including policies aimed at reducing the consumption of discretionary table salt.

## Introduction

Non-communicable diseases (NCDs) are a global health problem and unhealthy diets are a leading driver. Among dietary risk factors, high sodium intake was the leading cause of morbidity and mortality worldwide, accounting for some 3 million deaths and 70 million DALYs (1). In Brazil, NCDs are responsible for 75% of all deaths and CVD is the most frequent cause of death among NCDs (2). Cardiovascular disease (CVD) represents the biggest disease burden attributable to high sodium intake, and much of this risk is mediated through blood pressure increase (3).

The average daily sodium consumption in Brazil is approximately double the World Health Organization (WHO 2013) recommended maximum limit of 2g (4). Thus, over 27 thousand deaths from coronary heart disease and stroke could be prevented or postponed every year if Brazilians consumed, in average, less than 2 g/day sodium (5). However, unlike many high-income countries with ‘Western’ type diets, only about 35% of Brazilians’ dietary sodium comes from industrialized foods and salt-based condiments, whereas over 55% comes from added table salt (6)(7)(8). Sodium reduction policies in Brazil have therefore incorporated a set of strategies, These include health education campaigns targeted to the table salt added to foods and meals (9) and, since 2011, food reformulation strategies aimed at reducing the sodium content of processed and ultra-processed products, including condiments, through voluntary upper limits for sodium content in priority food categories (10). From 2011 to 2017, all food categories have reduced their upper limits of sodium and most have reduced their average sodium content, from 8% to 34% (11).

The objective of this study was to quantify the potential health and economic impacts of the voluntary sodium reformulation in Brazil.

## Methods

We have developed IMPACT _NCD BR_, a new microsimulation for Brazil using available local data, building on our previous experience on sodium modelling (12)(13)(14)(15). We used IMPACT _*NCD BR*_ to assess the potential health and economic effects of the voluntary targets for sodium content in processed foods in Brazil over a 20-year period (2013-2032).

We simulated the long-term impact of the reduction in the sodium content of processed and ultra-processed foods through the national voluntary targets from 2013 to 2017, compared to a “no intervention” baseline scenario. We assumed that only industries that officially committed to the voluntary targets would reduce the sodium content of their products and that the sodium content and target compliance by industries from 2017 onwards (11) would not change.

### The IMPACT _NCD-BR_ Model

IMPACT _NCD-BR_ is a stochastic, dynamic, microsimulation model based on the simulated adult life course of a close-to-reality synthetic individuals under different policy scenarios, considering the population heterogeneity and the lag times between exposures and outcomes. The model inputs, structure, key assumptions are detailed in S1 Appendix.

The model is designed to first run the “no intervention” scenario, simulating the life courses of the synthetic individuals and recording sodium consumption, systolic blood pressure, first cardiovascular episode and death (from CVD or any other cause). Then it simulates the life courses of the same synthetic population under the intervention scenario (in this case, food reformulation through the national voluntary sodium targets) and records its outcomes.

The close-to-reality population built in the model mirrors the Brazilian national population structure by age and sex (16) and includes data from the National Health Survey (*Pesquisa Nacional de Saúde -* PNS 2013)(17) and the National Household Budget Survey (*Pesquisa de Orçamentos Familiares -* POF 2008-2009)(18)(19) regarding sodium and systolic blood pressure (SBP) exposure.

### Microsimulation Model Structure

Figure 1 shows a simplified structure of the IMPACT _NCD-BR_ model. The model tracks individual□level sodium consumption, considering the different sources of dietary sodium, its impact on SBP, and the consequent risk of developing CHD, stroke, and death from these or any other cause. IMPACT _NCD-BR_ is calibrated to forecasts of CHD, stroke, and any□other□cause mortality for the whole Brazilian population from 2013 to 2032. The results are presented for adults aged 30 to 79 years within the simulation horizon of 20 years.

**Figure 1.**
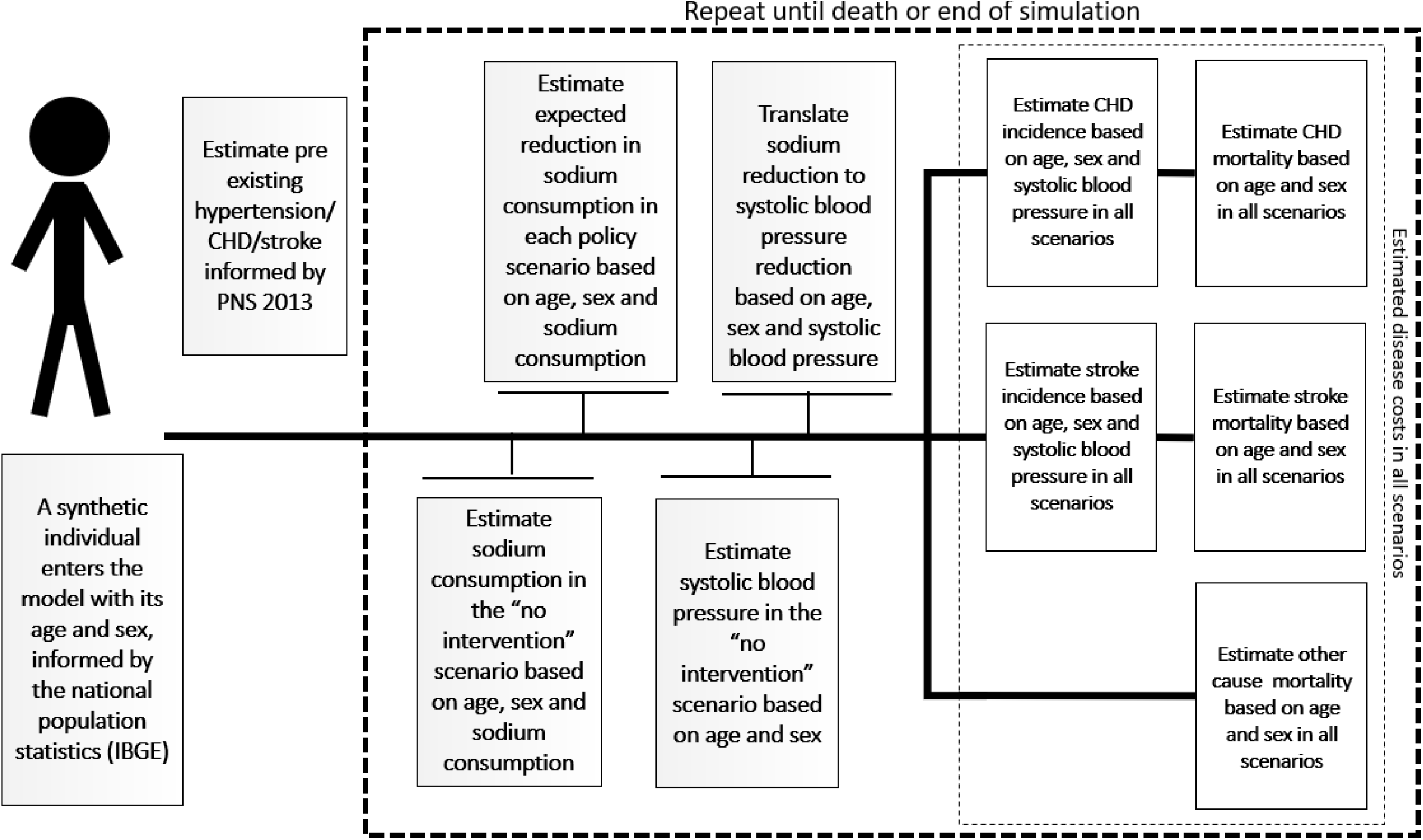
Simplified Structure of the Brazil IMPACT Sodium Policy Model.

For this, the model draws the traits of the synthetic individuals from conditional distributions, projects the sodium intake into the future and uses the projections to evolve the traits of the synthetic individuals over time. We used PNS 2013 for the SBP projections (assuming SBP remains constant over time for all age and sex groups) and POF 2008-2009 for the sodium intake projections (IBGE, 2014)(IBGE, 2010),(IBGE, 2011).

A more detailed model description and validation is provided in the S1 Appendix.

### Summary of evidence regarding the risks of excess sodium consumption

Excess dietary sodium consumption has been linked to an increased risk of CVD (20). For CVD, the excess risk appears to be mainly mediated through the deleterious effect of excess sodium consumption on blood pressure (BP) (3). Our methods for evaluating the causality of effects of sodium reduction on BP, and of BP reduction on CVD have been previously described (3).

### Policy effects

Brazil’s Ministry of Health has set voluntary reduction targets with food industries, from 2011 to 2017, based on upper bound sodium concentration targets for 34 food categories (10)(21). In addition, publicly available data from the Household Budget Survey (POF 2008-2009) was used to map these 34 food categories through household food acquisition (18) and a 24-hour recall dietary questionnaire (19). These data enabled the model to estimate the potential impact of the modeled policies on every synthetic individual based on their age and sex, and sodium consumption in the “no intervention” scenario. The model then used the estimated reduction in sodium consumption of the synthetic individuals to calculate the effect upon their SBP using a published meta-regression equation (3).

We assumed that reformulation of food products would adjust sodium content to targets until 2017, and that this would lead to an immediate change in sodium intake in synthetic individuals according to the level of reformulation. We also assumed that the reformulated products would thereafter sustain their sodium concentration.

Although changes in sodium intake influence SBP within weeks (3)(22), we conservatively assumed a median duration of 5 years from change in SBP to the health outcomes of CVD cases and CVD deaths.

### Modeling of food composition and sodium intake changes

We have considered changes in food composition from the voluntary sodium targets using data from official national food labeling surveys in 2013 and 2017: the baseline targets, and the most recent documented official monitoring respectively (11). Changes in sodium intake were modeled using microdata from the 2002-2003 and 2008-2009 Brazilian Household Budget Survey. We assumed that the average food consumption and use of table salt in the population remained constant from 2011 to 2017, and that sodium content was reduced for the targeted food categories only by industries that have voluntarily committed to the national sodium targets.

We used the publicly available data from the Household Budget Surveys (POF) of 2002-2003 (23) and 2008-2009 (18) in order to estimate the changes in sources of dietary sodium (non-industrial and industrial sources) during the period between both surveys, and projected the continuity of the replacement of discretionary salt (table salt) by processed and ultra-processed foods (representing the other sodium sources), assuming that the replacement would continue at the same rate in the future.

### Model outputs

For each scenario, the model generated the total numbers of relevant events and reported cases and deaths prevented or postponed (CHD or stroke or CVD or other). We present the results for Brazilian adults aged 30 to 79 years from 2011 to 2032 (simulation horizon of 20 years), rounded to 2 significant digits.

### Medical costs analyses

The CHD and stroke hospitalization costs to the Brazilian National Health System (SUS – *Sistema Único de Saúde)* were obtained from the publicly available tables from the Hospital Information System - SIH-SUS (24). For costs without readily available Brazil-specific data, we estimated the primary health, outpatient, pharmaceutical and informal care costs using the ratios of these costs to hospital activity costs from a study in European countries (25). Informal care costs in this study were applied based on the proportion of individuals with CVD who were hampered in daily activities. The average hours of informal care was then multiplied by average hourly wages for employed, working age carers, and minimum or lowest sectoral hourly wage for retired or not employed carers. Cost savings to the health system and to the population were estimated considering the cases of CHD and stroke prevented or postponed, and the average costs for a person-year living with disease. Costs were collected in Brazilian Reals (R$) and subsequently converted to U.S. dollars (US$), at an exchange rate of US$1 = R$3.876, current as of December 31, 2018, as reported by the Central Bank of Brazil.

### Sensitivity analyses and uncertainty analyses

We performed probabilistic sensitivity analyses via a second-order Monte Carlo approach for estimating the uncertainty of different model parameters and population heterogeneity to be propagated to the outputs (26). Uncertainty was based on sources of the sampling errors of baseline sodium intake, baseline SBP, and the relative risk of CHD and stroke based on SBP, the uncertainties around the lowest sodium and SBP exposures below which no risk is observed, around the effect of sodium on SBP, and around the true incidence of CHD and stroke, the uncertainty of mortality forecasts, and the uncertainty of all the costs. Output distributions were summarized by reporting the medians and 95% uncertainty intervals (UIs).

## Results

### Health related outcomes

The modelled results suggest that the recent voluntary sodium reduction policies in Brazil have led to a 0.249 g/day decline in salt consumption (0.1 g/day of sodium), from 2011 to 2017. If that 0.1g/day sodium reduction continues, it could potentially prevent or postpone approximately 2,500 CVD deaths (95% UI: −900 to 11,000) and some 182,000 CVD cases (95% UI: 45,000 to 425,000), as well as 11,500 deaths from other hypertension-related causes (95% UI: 3,600 to 22,000), from 2013 to 2032 (Table 2).

**Table 2.** Health-related model estimates over a 20-year simulation period, from 2013 to 2032, for Brazilian adults aged over 30 years.

| **Outcome** | **Men** | **Women** | **Total** |
| --- | --- | --- | --- |
| CHD cases prevented or postponed | 67,000<br>(17,000 to 157,000) | 31,000<br>(6,000 to 78,000) | 98,000<br>(23,000 to 235,000) |
| Stroke cases prevented or postponed | 45,000<br>(11,000 to 101,000) | 39,000<br>(10,000 to 89,000) | 84,000<br>(22,000 to 190,000) |
| CHD deaths prevented or postponed | 700<br>(-200 to 900) | 500<br>(-200 to 2,000) | 1,200<br>(-500 to 5,000) |
| Stroke deaths prevented or postponed | 700<br>(-200 to 3,000) | 700<br>(-200 to 2,800) | 1,400<br>(-500 to 6,000) |
| Non-CVD deaths prevented or postponed | 6,900<br>(2,300 to 12,800) | 4,600<br>(1,400 to 9,200) | 11,500<br>(3,700 to 22,000) |
| All deaths prevented or postponed | 8,300<br>(1,800 to 19,000) | 5,800<br>(900 to 14,000) | 14,000<br>(2,700 to 33,000) |

**Table 3.**
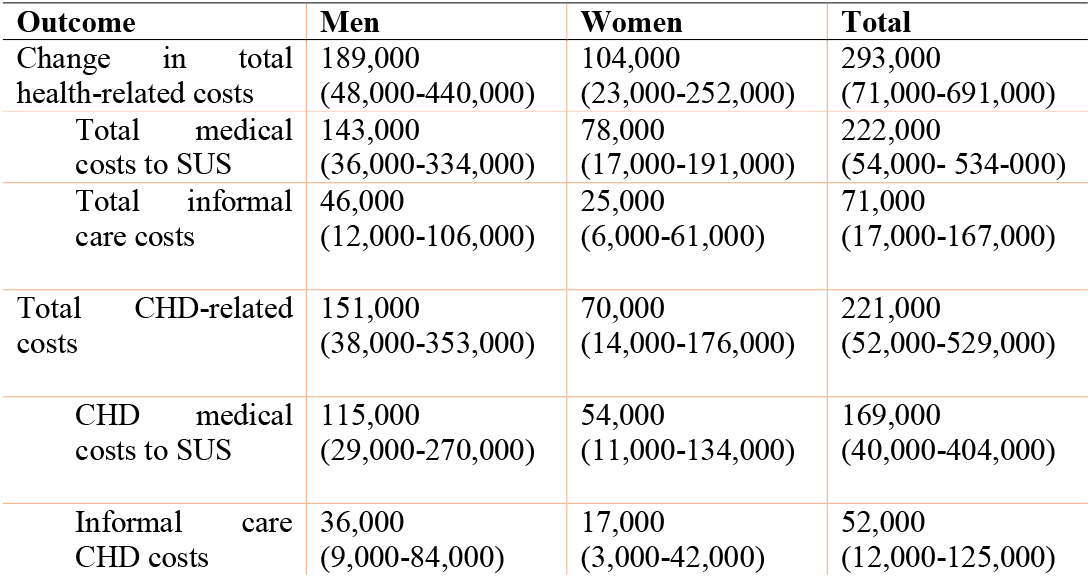
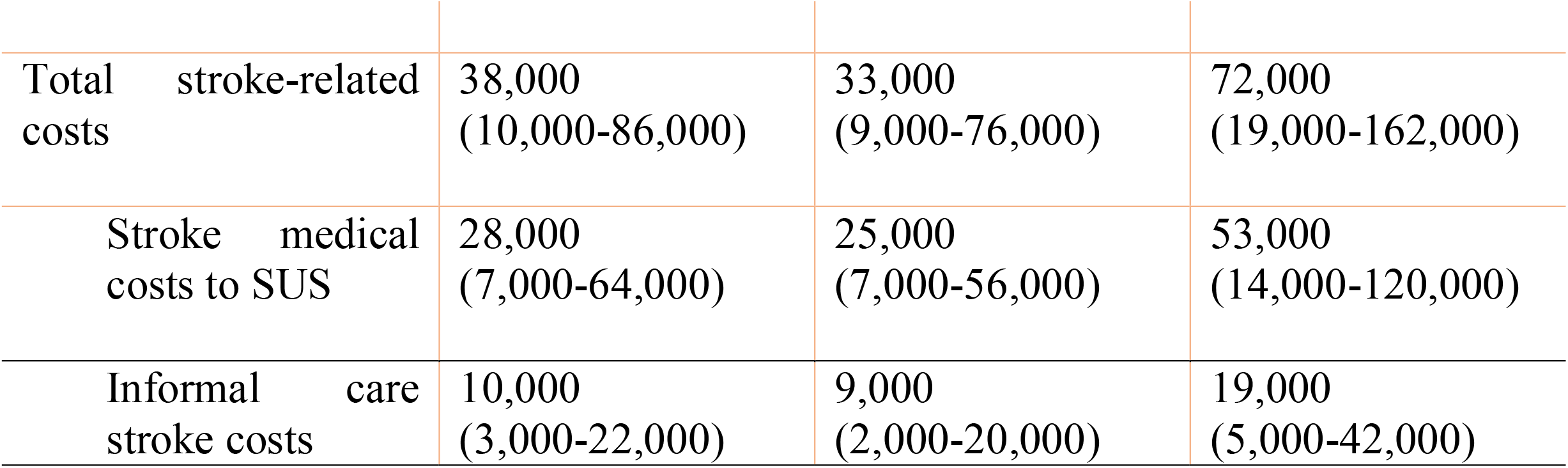
Impact inventory and cost analysis of CVD-related model outputs for individuals aged 30 to 84 years, assessed cumulatively over the 20-year simulation period from 2012 to 2032 (US$ thousands).

The health benefits of sodium reduction, especially in reducing coronary heart disease, were larger in men than women, reflecting men’s higher sodium consumption and higher CVD burden. In total, during the 20-year time period from 2013-2032, the voluntary sodium targets could prevent or postpone approximately 112,000 CVD cases (95% UI: 28,000 to 258,000) in men and some 70,000 cases in women (95% UI: 16,000 to 167,000), and approximately 1,400 fewer CVD deaths in men (95% UI: −500 to 4,100) and 1,200 fewer deaths in women (−500 to 4,800,).

Almost two thirds of the CVD cases and CVD deaths that would be prevented or postponed by this policy between 2013 and 2032 would be in people aged 50 to 69 years (Appendix).

### Costs of CHD and stroke

From the public healthcare perspective, the voluntary targets for sodium could result in a net saving of approximately US$ 290 million (95% UI: US$ 70.7-691.3 million) in cumulative hospitalization, primary health, outpatient, pharmaceutical, and informal care costs, over the 20-year period. Most of the savings would be related to coronary heart disease (75%) rather than stroke.

The US$ 290 million estimated savings through the continuity of the voluntary sodium targets would come from

a. Approximately US$ 220 million savings (95% UI: US$ 53.6-524.4 million) related to reduced CHD and stroke treatment costs to the Brazilian National Health System.
b. Approximately US$ 70 million (95% UI: US$ 17.1-166.9 million) savings in informal care costs.

## Discussion

We have identified potentially large future health and economic benefits if Brazil were to continue the voluntary sodium targets first set in 2011. We used a previously validated microsimulation approach to create a close-to-reality synthetic population (IMPACT_NCD-BR_ Model) for the Brazilian population. Our analysis suggests that continuing the voluntary targets could result in substantial decreases in CVD incidence and mortality whilst also offering large cost savings to the public healthcare system and individuals.

Our findings highlight the substantial health and economic opportunity costs of inaction and that, despite contributing to reduce the burden of CVD, the voluntary targets on processed and ultra processed foods need to be more stringent and to be accompanied by other strategies in order to promote more significant sodium reduction in the Brazilian population through other dietary sources.

The estimated 0.1g reduction in daily sodium consumption from 2011 to 2017 achieved by the voluntary targets in Brazil has been modest when compared with some other countries. A systematic review of 70 papers concluded that multi-component strategies involving both upstream and downstream interventions, generally achieved the biggest reductions in salt consumption across an entire population, most notably 4g/day in Finland and Japan, 3g/day in Turkey and 1.3g/day recently in the UK (8). Mandatory reformulation alone could achieve a reduction averaging around 1.45g/day (three separate studies), followed by voluntary reformulation (−0.8g/day), school interventions (−0.7g/day), short-term dietary advice (−0.6g/day) and nutrition labelling (−0.4g/day). Tax and community based counselling could, each typically reduce salt intake by 0.3g/day, whilst even smaller population benefits were derived from health education media campaigns (−0.1g/day). Although long-term dietary advice could achieve a - 2g/day reduction under optimal research trial conditions; however, smaller reductions might be anticipated in unselected individuals (8). Furthermore, the burden of disease attributable to excessive dietary sodium remains large, almost 30 thousand avoidable coronary heart disease and stroke deaths every year (5).

The voluntary targets reduced the average salt consumption by 0.249 g/day (0.1 g/day of sodium) between 2013 and 2017 (a 25 mg/day reduction every year), a reduction of just 2%. By contrast, the United Kingdom achieved an average annual reduction of 0.08 g/d among men and by 0.05 g/d among women (13) between 2003 and 2010. However, in high income countries such as the

United Kingdom, some 80% of dietary sodium comes from processed and ultra-processed foods, compared with only about 35% in Brazil; whereas over 55% comes from added table salt (6)(18).

The Brazilian voluntary sodium targets have resulted in a gradual sodium reduction, in order to avoid noticeable changes in the taste of foods and allow industries to develop new technologies to reduce or replace sodium (27)(28). Gradual reductions are unlikely to result in compensatory behaviors as adding more table salt to foods or while cooking (29)(30), while large reductions in a short period of time may trigger rejection by consumers (31).

In 2018, the attributable costs of hypertension to the Brazilian National Health System reached approximately US$ 525 million/year (32) and, in 2013, it was estimated that some US$ 100 million/year in public hospitalizations could be saved if sodium consumption were reduced to 2 g/day in Brazil (33).

This study is focused solely measuring industrial salt. This will become increasingly important given the increasing contribution of ultra-processed foods in diets. Food reformulation and other regulatory strategies will therefore become essential for improving the food environment, by providing healthier food options (8)..

Furthermore, reducing future population salt intake in the Brazilian context will depend both on strengthening industry action on sodium, but also improving strategies to modify population behaviour in relation to discretionary (non-industrial) salt use. Thus, the Brazilian Dietary Guidelines emphasize the importance of unprocessed and minimally processed foods as the foundation of diets, as well as a conscious use of culinary ingredients, as salt.

### Strengths and limitations of this study

The IMPACT NCD microsimulation models have been extensively validated and replicated in different countries (12)(14). This is the IMPACT model adapted to a Latin American country and represents a big step forward for using models to inform policy in the region. The study’s strengths also include the use of representative population data and of good effect sizes from meta-analyses.

This study also has potential limitations. The model’s effect estimates are based on interventional and prospective observational studies, and may therefore retain possible biases and confounding factors. However, the etiological effects of dietary changes were estimated from meta-analyses with confirmatory validity analyses, including from randomized clinical trials. The estimated benefits from the model may be conservative and underestimate the full health and economic health gains from sodium reformulation, as (1) the counterfactual scenario assumed that the participation of processed and ultra-processed foods in sodium intake would not change into the future; (2) the model only evaluated diseases mediated through BP, while decreasing sodium consumption could have beneficial effects upon other health burdens such as gastric cancer (34); and (3) food reformulation by industries might additionally increase potassium intake though substitution of NaCl with KCl (35), which potential beneficial effect was not included in the model. Finally, medical costs from the private sector, which covers 30% of the Brazilian population, were not included in the economic estimates, which will therefore be conservative.

### Policy implications

Our findings suggest that the voluntary sodium reduction targets for processed and ultra-processed foods in Brazil could generate health benefits to the population and cost savings to the National Health System and in terms of informal health treatment cost. Nevertheless, in order to achieve more substantial sodium reductions and its consequent health and economic impacts, lower sodium targets must be implemented across all food industries. Other dietary sources of sodium must also be tackled, especially through food-based dietary guidelines and comprehensive approaches to healthy diets. The potential health and economic benefits could be substantial.

## Data Availability

All data used in this manuscript was obtained from publically available databases and meta-analyses from literature

