## Supplementary materials for "ESTIMATING THE HEALTH AND ECONOMIC EFFECTS OF THE VOLUNTARY SODIUM REDUCTION TARGETS IN BRAZIL: MICROSIMULATION ANALYSIS"

The authors have provided this Appendix to give readers additional information about their work.

This Technical Appendix was adapted from a previously published one in:

Pearson-Stuttard J, Kypridemos C, Collins B, Mozaffarian D, Huang Y, Bandosz P, et al. Estimating the health and economic effects of the proposed US Food and Drug Administration voluntary sodium reformulation: Microsimulation cost-effectiveness analysis. PLOS Med. 2018;15:e1002551.

### Summary of evidence about the risks of excess sodium consumption

The body of evidence, observational and interventional, on the causal relationship between salt/sodium intake and blood pressure (BP) is large and expanding (1) (2). Increased intake of dietary sodium is associated with an increase in the risk of cardiovascular disease (3), mainly mediated through the deleterious effect of sodium on systolic blood pressure (SBP) (4) . In our model, we have explicitly modelled the causal pathways of sodium reduction on SBP and of SBP reduction on CVD risk.

The World Health Organization (WHO) guidelines recommend a daily sodium intake of less than 2,000 mg/d (5 g/d of salt), and national recommendations in Brazil have followed (5). Despite recent studies that have questioned the optimum levels of salt/sodium consumption recommended to populations (6)(7)(8), strong evidence has supported that inaccurate methodologies bias these controversial studies (1)(9)(10). Therefore, current salt intake recommendations are based on robust scientific evidence and are vital to promoting health across the world (11)(12)(13)(14). A meta-analysis of studies from different countries has concluded that the optimal level of sodium consumption below which no health gains have been observed is somewhere in the range of 614 mg/d to 2391 mg/d (2), so we have incorporated the uncertainty around the ideal sodium consumption in our probabilistic sensitivity analysis.

Strong and abundant existing evidence supports that the effect of low sodium diet on blood pressure appears to happen within weeks (15) and the cardiovascular risk reversibility of blood pressure appears to occur within five years, according to several randomised control trials (16).

### A high-level description of the IMPACT _NCD BR_ model

The IMPACT _NCD BR_ model is a discrete-time dynamic stochastic microsimulation model (17). It is an implementation of the IMPACT_NCD_ modelling framework that has been used previously to model the impact of sodium reduction policies in England and the US (18)(19)(20).

Within the IMPACT _NCD BR_ model, each unit is a synthetic individual and is represented by a record containing a unique identifier and a set of associated attributes. For this study, we considered age, sex, sodium consumption (considering industrial and non-industrial sources of dietary sodium, i.e. the sodium added by industries to foods and the other sources of sodium in the diet, including table salt), and SBP. A set of stochastic rules is then applied to these individuals, such as the probability of developing coronary heart disease (CHD) or dying, as the simulation advances in discrete annual steps. The output is an estimate of the burden of CHD and stroke, in the synthetic population, including both total aggregate change and, more importantly, the distributional nature of the change (Figure 1).

The IMPACT _NCD BR_ model is a complex model that simulates the life course of synthetic individuals and consists of four simulation engines: the 'population' engine, the 'disease' engine, the ‘health economics’ engine, and the 'policy' engine. The description of the processes in each of the engines is fully described in the following chapters. The description is focused on the model's rationale from an epidemiological perspective and complements the source code that is available at Github (https://github.com/ChristK/IMPACTncd_Br/tree/voluntary_reformulation) covered by the GPL v3 licence. Figure S1 depicts the logic of the model and Tables A and B summarise the sources of the input parameters and the main assumptions and limitations, respectively.

Technical information

The IMPACT _NCD BR_ model is being developed in R v3.6.1. The IMPACT _NCD BR_ model is built around the R package 'data.table', which imports a new heavily optimised data structure in R. Most functions that operate on a data table have been coded in C to improve performance. To ensure statistical independence of the pseudo-random number generators running in parallel, the R package 'doRNG' was used to produce independent random streams of numbers, generated by L'Ecuyer's combined multiple-recursive generator.

Figure 1. Logical framework for the IMPACT _NCD BR_ model

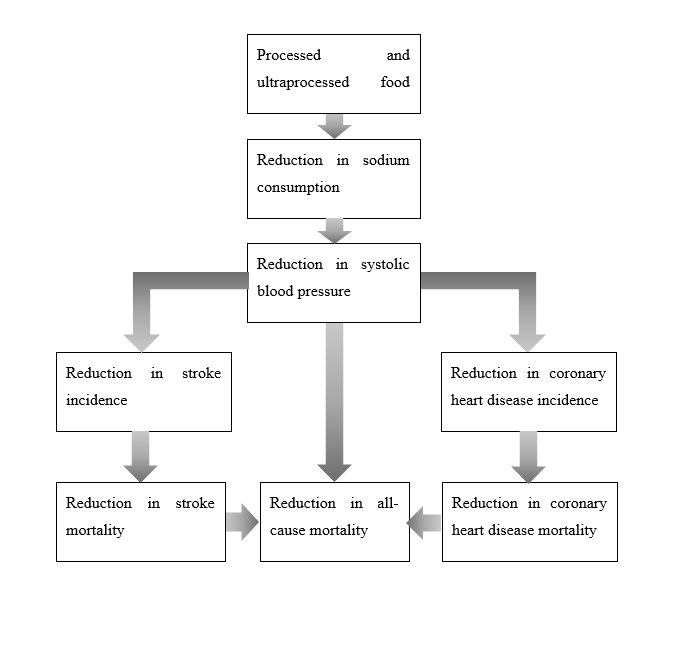

### Population engine

The population module includes the demographic module and the exposure module.

#### Demographic module

Synthetic individuals enter the simulation in the initial simulation year (2013 for this study). The number of synthetic individuals that enter the simulation is user-defined, and for this study was set to 400,000. The algorithm ensures that the joint age and sex distribution of the sample are as this of the Brazilian population in 2013 (21). Then it creates backward and forward projections of the synthetic population that are essential to model exposure time trends and time lags between exposures and diseases.

The backward projection of the synthetic population goes back to 2003; therefore, the maximum time lag we allow in the model is ten years. As everyone alive and older than 30 years old in 2013 was alive in 2003, the algorithm simply creates the back projections by appropriately reducing the age of the synthetic individuals, while keeping constant all other variables.

For the forward projections, we project until the year 2032, and the algorithm increases the age of the synthetic individuals while keeping the sex variable constant. For forward projections, mortality needs to be considered. We describe mortality with the disease module as disease-specific mortality which is closely related to disease prevalence. The model follows an open cohort approach. Every simulated year from 2013 onwards, a new cohort of 30 years old synthetic individuals enters the model. The same sources inform the size of the cohort and the joint age and sex distribution we described above. For example, in 2014, the new 30-year old cohort will be informed by the population size and the joint age- sex -distribution of those who were 29 years old in 2013. The approach may be crude; however, the final model outputs are directly standardised to Brazilian population projection (21) to minimise the bias.

#### Exposure module

This module simulates the adult life course sodium and SBP exposures of the synthetic individuals based on POF 2008-2009^[1]^ the National Health Survey 2013 (PNS 2013) (22). For all simulated exposures, we followed the same general principles. First, we fitted a Generalised Additive Model for Location, Scale and Shape (GAMLSS)^[2]^ to the data with the exposure of interest as the dependent variable, and some functions of age and sex as independent variables (23). Then, we use the GAMLSS model to predict the exposure level of every synthetic individual in the simulation and simulate individualised risk factor trajectories for all synthetic individuals, based on their age and sex that were estimated from the sociodemographic module.

The approach described above provides us with equations to estimate the distribution of an exposure for a given age and sex. When the synthetic individual enters the simulation, a set of random numbers between 0 and 1 and of size equal to the number of the modelled exposures is allocated to her. Each one of them represents the percentile of the relevant exposure distribution. The principle is that synthetic individuals retain their percentiles throughout their life course (this is known as the rank stability assumption) (24). For example, in 2013, a 40-year-old male synthetic individual with SBP of 120 mmHg has a SBP percentile of 0.52. Twenty years later, the same synthetic individual has retained his percentile score for SBP. However, his SBP is now estimated to 137.6 mmHg because the SBP distribution has changed to reflect the SBP of 60-year old men.

Finally, to model the exposure to sodium intake we fitted to GAMLSS models, one for industrial and one for non-industrial sources of dietary sodium. Because these two exposures were correlated, we simulated their linear correlation by using correlated random number streams for the percentiles.

### Disease engine

The previous two modules for demographics and exposure generate a dynamic close-to-reality synthetic population that is composed of the adult life course exposures of each of the synthetic individuals. The disease module then translates these exposures to disease incidence, using a population attributable risk approach (PAF) (25). We will first describe how disease incidence is simulated in the model, and then how the model simulates mortality.

#### Disease incidence

To estimate the individualised annual probability of a synthetic individual developing a specific disease conditional on their risk exposures, we follow a 3-step approach:

Step 1. The proportion of incidence attributable to SBP by age and sex is estimated, assuming a mean time lag of 5 years between exposure and disease, reflecting the best possible empirical data based on the observation period of cohort studies and time to risk reversal in randomised clinical trials (16)(26)(27).^[3]^ In each iteration the time lag varies stochastically between 2 and 10 years following a shifted binomial distribution.

Step 2. The portion of the disease incidence attributable to SBP is estimated and subtracted from the total incidence for 2013.

Step 3. The probability of developing the disease is estimated for each individual in the synthetic population and is used in an independent Bernoulli trial to select those who finally develop the disease.

The implementation of the above method is described in more detail using CHD as an example. The same process is used for stroke.

Step 1

PAF is an epidemiological measure that estimates the proportion of the disease attributable to an associated risk factor. It depends on the relative risk associated with the risk factor and the prevalence of the risk factor in the population. In a microsimulation context where exposure to risk factors are known at the individual level, PAF can be calculated using the formula:

$$PAF= 1-\frac{n}{\sum_{i=1}^{n} RR_{i}}$$

where $n$ is the number of synthetic individuals in the population, and ${RR}_{i}$ are the relative risks of the SBP associated with CHD, for each individual $i$. We calculated PAF based on above formula stratified by age and sex only in the initial year of the simulation. Consistent with findings from the respective meta-analyses that were used for the IMPACT _NCD BR_ model (Table A), SBP below 110 mmHg, was considered to have a relative risk of 1. All the relative risks were taken from published meta-analyses (Table A).

Step 2

The incidence of CHD not attributable to the modelled risk factors can be estimated by the formula:

$$I_{Theoretical minimum}= I_{Observed}*\left( 1-PAF \right)$$

Where $I_{Observed}$ is the CHD incidence and $PAF$ is from Step 1. $I_{Theoretical minimum}$ represents CHD incidence if SBP was at optimal levels across the population. To account for future time trend in CHD incidence that is not attributable to SBP, the model updates $I_{Observed}$ every simulated year. For this we assume that half of the forecasted annual change in CHD mortality is attributed to changes in CHD incidence and the other half to changes in CHD case fatality. We based this assumption on observational evidence from England, and modeling studies in the England and the US (28)(29)(30)(31). Furthermore, we included this assumption in our probabilistic sensitivity analysis.

Stage 3

Assuming that $I_{Theoretical minimum}$ is the annual baseline probability of a synthetic individual to develop CHD for a given age and sex due to risk factors not included in the model, the individualised annual probability of developing CHD, $\mathbb{P}\left( \text{CHD | age, sex, exposures} \right)$, given his/her risk factors were estimated by the formula:

$$\mathbb{P}\left( CHD | age,sex, exposures \right)= I_{Theoretical minimum}*RR_{i}$$

Where $RR_{i}$ the relative risk that is related to the SBP of the synthetic individual, same as in stage 1.

The method described above can be used only when the incidence of the disease in the population is known. However, the true incidence of CHD (and stroke) is mostly unknown. While several estimates exist, all have limitations. Therefore, for the estimation of CHD incidence by age and sex, we opted for a modelling solution to synthesise all the available sources of information and minimise bias. Specifically, we used the Information System on Mortality (*Sistema de Informações de Mortalidade* - SIM) database (32) to extract mortality rates for CHD (ICD-10: I20–I25) for the years 1999–2015, stratified by age and sex. We additionally estimated self-reported prevalence of CHD by age and sex from the PNS 2013 (22). We used both prevalence and mortality rates to inform the World Health Organisation (WHO) DISMOD II model (33). DISMOD II is a multi-state life table model that can estimate the incidence, prevalence, mortality, fatality, and remission of a disease when information about at least three of these indicators is available. A similar approach has been followed by the Global Burden of Disease team and other groups (34)(35). We considered CHD an incurable chronic disease (i.e. remission rate was set to 0); therefore, the derived DISMOD II incidence refers to the first-ever manifestation of angina or AMI excluding any recurrent episodes. For the DISMOD II calculations, we assumed that incidence and case-fatality had each been declining by 2% (relative), over the last 20 years. The derived CHD incidence and prevalence rates were used as an input for IMPACT _NCD BR_. A similar appr(35)oach was used for stroke.

For the initial simulation year, some synthetic individuals need to be allocated as prevalent cases for each of the modelled diseases. We use DISMOD II prevalence estimates to identify prevalent disease cases by age, sex.

#### Mortality

All synthetic individuals are exposed to the risk of dying from any of their acquired modeled diseases or any other non-modeled cause in a competing risk framework. The IMPACT _NCD BR_ model is calibrated to observed CHD, stroke, and any-other-cause mortality for year 2013 (IBGE, 2014) and mortality forecasts for years 2013–2032. For years after 2017, coherent functional demographic models by sex and age were fitted to the reported CHD, stroke, and any-other-cause mortality rates from years 2000 to 2017 (32), and then were projected to the simulation horizon using the R package 'demography' (36). Functional demographic models are generalisations of the Lee-Carter demographic model, influenced by ideas from functional data analysis and non-parametric smoothing (37). The coherent approach ensures that subgroup forecasts do not diverge over time (38). Finally, we used the observed and forecasted mortality rates to create life tables for each simulated year, by age, sex, and disease (CHD, stroke, any-other-cause). We applied the any-other-cause life tables to all synthetic individuals, and the CHD and stroke life tables to prevalent cases of CHD and stroke only, respectively. For the synthetic individual that died of more than one causes in a specific year, a cause was randomly selected to minimise bias.

In reality, hypertensive individuals have a higher risk to die not only of CHD and stroke but from a spectrum of other diseases also. To account for this and minimise bias, the IMPACT _NCD BR_ model inflates the any-other-cause mortality rates for hypertensive synthetic individuals in the model (Figure A link between SBP and all-cause mortality) while it deflates it for non-hypertensives. The algorithm ensures the total number of hypertensive and non-hypertensive synthetic individuals that die every year from any-other-cause is equal to the defined one in the life table. The algorithm is based on PAF approach, and the relative risk was derived from an individual level meta-analysis by Stringhini *et al* (39). In this meta-analysis the relative risk of all-cause mortality for hypertensives was 1.31 (1.24–1.38), and the relative risk of non-CVD-non-cancer mortality was 1.29 (1.21–1.38). Hence, we used a relative risk of 1.3 in the IMPACT _NCD BR_ model.

### Health economics engine

In the previous two modules, the IMPACT _NCD BR_ model creates synthetic individuals with traits similar to those observed in the Brazilian population and tracks their future exposures to sodium and SBP, and important events (first manifestation of CHD and stroke, death from CHD, stroke, or any other cause).

#### Disease costs

The IMPACT _NCD BR_ model applies CHD, stroke, and hypertension costs to cases of these diseases, during the simulation. These costs are mean estimates by age and sex.

Disease costs per person-year were derived from the National Health System's Hospital Information System (SIH/SUS) (40). We assumed constant medical costs in US dollars. Medical costs per person-year for CHD and stroke were calculated by dividing total hospitalisation costs by the number of people with each condition in 2017.

Informal care costs for CHD and stroke were based on the ratio of hospital care to other medical costs in Europe from a study by Leal *et al.* (41). We assumed no informal care costs for hypertension alone.

### Policy engine

Until now, the description of the IMPACT _NCD BR_ model was for the baseline scenario. The policy module translates the policy scenarios to be modeled by the IMPACT _NCD BR_ model. Changes in sodium consumption are translated into changes in SBP using the meta-regression equation by Mozaffarian *et al*. (2) by age and hypertensive status.^[4]^ The new SBP is used in the disease module, and updated CHD and stroke risks are calculated for every synthetic individual, with new outcomes. Therefore, new life courses for all synthetic individuals are simulated as a result of the modeled policies. At the end of the simulation, the model compares all alternative life courses with the baseline life course for each synthetic individual and calculates the outputs.

#### Modeling the proposed Brazilian voluntary sodium targets

In 2010, the Brazilian Ministry of Health proposed a voluntary approach to reduce the average sodium content of the industrialised foods which contributed to over 90% of the sodium intake from processed and ultraprocessed foods, partnered by the Brazilian Association of Food Industries (Abia) (42). The targets were set as gradually decreasing upper limits to salt content in foods, in two-year steps. Because target setting was negotiated individually for each food category, the agreements (Terms of Commitment) were released gradually, from 2011 to 2013 (43)(44)(45)(46), until all priority food categories had targets. The targets were monitored through nutritional label surveys in 2013-2014 and 2017-2018 (47)(48)(49), which analysed the compliment to the targets and the reduction in the average sodium content of foods during the period (50)(51).

Separately, we linked the priority food categories to the POF 2008-2009 foods and codes in order to re-estimate sodium intake at baseline and in 2017. Therefore, to model the effect of the proposed policy to the modeled population we developed the algorithm below:

##### Step 1

We used the POF 2002-2003 and 2008-2009 food acquisition data to estimate the annual change in the participation of sodium sources in diet (added salt to foods and existing sodium in processed and ultraprocessed foods) and assume that the linear change in the dietary sodium sources would continue until 2032. Then, the distribution of sodium intake from the POF 2008-2009 24h Food Recalls was adjusted according to the estimates from the food acquisition module and the average sodium intake by age-group and sex were recalculated.

##### Step 2

We considered an average market share of 70% for all products from industries associated to the Brazilian Associations of Food Industries (Abia) and used the food acquisition microdata from the POF 2008-2009 survey. The sodium baseline content of targeted foods was replaced by the average content of these foods at the beginning of target negotiations and was recalculated for 2017 according to the estimated changes in the monitoring results (51). As applied by Sarno et al (52), the final sodium intake was adjusted to a 2,000 kcal/day diet in order to estimate the contribution of foods consumed out of the household.

##### Step 3

Finally, we apply the expected relative sodium reduction to the baseline sodium consumption of the synthetic individuals to estimate the net effect of the policy. This net policy effect on every synthetic individual, expressed in a change in sodium consumption every year, is transformed to SBP changes as it was described above. The underlying assumptions in this step is that total sodium consumption remains similar to 2013, that the total consumption of each priority food category remains similar to that in 2008-2009, and that the sodium content of other foods would remain unchanged.

This approach allows the incorporation of sodium consumption time trends in the calculations and provides enough granularity of the policy effect (by age, sex, and sodium consumption) without being too computationally intensive. Yet, it does not address potential behavioral changes of the population as a result of the reformulation and does not account for foods prepared in food outlets and restaurants.

### Uncertainty and sensitivity analysis

The IMPACT NCD BR model implements a 2^nd^ order Monte Carlo approach to estimate uncertainty intervals (UI) for each scenario (53). Each simulation, which includes all policy scenarios, runs 2000 times. For each iteration, a different set of input parameters is used by sampling from the respective distributions^[5]^ of input parameters, and a different sample of 400,000 synthetic individuals is drawn. Then, the life course of every synthetic individual is simulated for the baseline, and all policy scenarios and the outcomes are collected and summarised for the population, annually.^[6]^ Therefore, all model outputs (cases and deaths prevented or postponed) are separately estimated for each iteration, and conditional on the set of model inputs.

The framework allows stochastic uncertainty, parameter uncertainty, and individual heterogeneity to be reflected in the reported UI. The following example illustrates the different types of uncertainty that were considered in the IMPACT _NCD BR_ model. Let us assume that the annual risk of CHD is 5%. If we apply this risk to all individuals and randomly draw from a Bernoulli distribution with $p$ = 5% to select those who will manifest CHD, we only consider stochastic uncertainty. If we allow the annual risk for CHD to be conditional on individual characteristics (i.e. age, sex, exposure to risk factors), then individual heterogeneity is considered. Finally, when the uncertainty of the relative risks due to sampling errors is considered in the estimation of the annual risk for CHD, the parameter uncertainty is considered. From these three types of uncertainty, only the parameter uncertainty can be reduced from better studies in the future.

The structure of the model is grounded on fundamental epidemiological ideas and well-established causal pathways; therefore, we considered this type of uncertainty relatively small and did not study it. However, the discrete-time nature of the model can potentially introduce bias in cases where the synthetic individual dies more than once within a year, and the model cannot identify which event happened first. As we describe earlier, to minimise this type of bias we randomly select one of the events to be considered as it happened before all others, whenever these cases arise during the simulation.

#### Input uncertainty

The sources of uncertainty we considered were:

1. The sampling error of the baseline sodium intake.
2. The sampling error of the baseline SBP.
3. *The sampling error of the relative risks of SBP on CHD, stroke, and any-other-cause mortality*. We used the reported relative risks and their confidence intervals to construct log-normal (uniform for *any-other-cause mortality*) distributions (Table A).
4. The uncertainty around the lowest exposure to sodium below which no risk is observed. We used evidence in Mozaffarian et al. as parameters for a Pert distribution (Table A).
5. The uncertainty around the lowest exposure to SBP below which no risk is observed. We used evidence in Singh et al. (Table A).
6. *The uncertainty around the effect of sodium on SBP*. We used the meta-regression equation in Mozaffarian *et al*. Each time the model uses the equation a new set of coefficients was sampled from their respective normal distributions (Table A).
7. *The uncertainty around the lag time of SBP exposure and disease outcomes.* The distribution 1 + Binomial(9, (5-1)/9) to vary lag time between 1 and 10 years (median 5 years).
8. *The uncertainty around the true incidence and prevalence rates of CHD and stroke*. We described in page 14 how we used DisMod II to estimate the incidence rate of CHD and stroke. We fitted beta distributions by age, sex, and race/ethnicity assuming the 0.025 percentile to be half of the central estimate, the median the central estimate, and the 0.975 percentile double the central estimate.
9. *The uncertainty of mortality forecasts*. We incorporated the predictive uncertainty of the mortality forecasts to the IMPACT _NCD BR_ model estimates.
10. The uncertainty around the assumption that half of the forecasted annual change in CHD and stroke mortality is attributed to changes in CHD and stroke incidence, respectively. We allowed this assumption to vary, independently for each disease, between 0% and 100% following a uniform distribution.

#### Outputs

We summarise the output distributions of the IMPACT _NCD BR_ by reporting the medians and 95% uncertainty intervals (UI). We also plotted the annual probability that a scenario was cost effective or cost saving over the simulation period. Table J presents model estimates for the baseline scenario.

*Cases (Deaths) prevented or postponed*, by comparing the life course of each specific individual in the baseline scenario with its life course in the policy scenario.

All outputs can be stratified by year, age, sex, and disease. Moreover, outputs are scaled to the Brazilian population (from the 400,000 sample of synthetic individuals).

It is important to not misinterpret 95% UIs as 95% confidence intervals (CI) and overlapping UIs as 'evidence against statistical significance.' This does not apply to our model outputs because the scenarios share common model inputs as explained above and should not be treated as 'independent' from a statistical perspective.

### Tables

Table A. The IMPACT NCD BR model data sources.

| Parameter | Outcome | Details | Comments | Source |
| --- | --- | --- | --- | --- |
| Population size estimates | Population | Resident population from 2010 National Census and intercensal estimates | Stratified by year, age, and sex | Brazilian Institute of Geography and Statistics (IBGE) – online reports tables and microdata [Internet]. 2017 (IBGE, 2011b)(IBGE, 2017a) |
| Population projections | Population | 2012–2060 Brazilian population projections produced by the Brazilian Institute of Geography and Statistics (IBGE) | Stratified by year, age, and sex | Brazilian Institute of Geography and Statistics (IBGE) – online reports and tables [Internet]. 2017 (IBGE, 2019) |
| Mortality | Deaths from CHD, stroke, and any other non-modeled causes | Underlying cause of death 2000-2017 | Stratified by year, age, sex, and cause of death | Ministry of Health of Brazil. National Mortality Information System (*Sistema de Informações de Mortalidade –* SIM). Underlying cause of death 2000-2016 based on the ICD-10 codes. 2018 (MINISTÉRIO DA SAÚDE, 2017a) |
| Exposure to sodium | Exposure of individuals | National Household Budgetary Surveys (POF) | Anonymized, individual-level data sets. Years 2008-2009. | Brazilian Institute of Geography and Statistics (IBGE) – POF 2008-2009 (*Pesquisa de Orçamentos Familiares*) online public microdata and reports [Internet]. 2011 (IBGE, 2011a)(IBGE, 2010) |
| Exposure to systolic blood pressure | Exposure of individuals | National Health Survey (PNS) | Anonymized, individual-level data sets. Year 2013. | Brazilian Institute of Geography and Statistics (IBGE) – PNS 2013 (*Pesquisa Nacional de Saúde*) online public microdata and reports [Internet]. 2014 (IBGE, 2014) |
| Effect of sodium consumption on systolic blood pressure | Systolic blood pressure change | Meta-analysis/meta- regression of 103 trials | Only trials with duration > 7 days were analysed. | Mozaffarian D, Fahimi S, Singh GM, *et al*. Global sodium consumption and death from cardiovascular causes. New England Journal of Medicine 2014;371(7):624–34. (Text S1 in the Appendix) (MOZAFFARIAN et al., 2014) |
| Setting reference level of sodium consumption | Ideal sodium consumption below which no risk was considered | Evidence from ecologic studies randomised trials and meta-analyses of prospective cohort studies | Intake levels associated with the lowest risk ranged from 614 to 2391 mg/day. In large, well-controlled, randomised feeding trials, the lowest tested intake for which blood pressure reductions were clearly documented was 1500 mg/day. | Mozaffarian D, Fahimi S, Singh GM, *et al*. Global sodium consumption and death from cardiovascular causes. New England Journal of Medicine 2014;371(7):624–34. (Text S4 in the Appendix and Table C) (MOZAFFARIAN et al., 2014) |
| Relative risk for systolic blood pressure | CHD and stroke (ICD10: I20–I25 and I60–I69) | Pooled analysis of two individual level meta-analysis | Stratified by age and sex. Adjusted for regression dilution and total blood cholesterol and, where available, lipid fractions (HDL and non-HDL cholesterol), diabetes, weight, alcohol consumption, and smoking at baseline. | Micha R, Peñalvo JL, Cudhea F, Imamura F, Rehm CD, Mozaffarian D. Association between dietary factors and mortality from heart disease, stroke, and type 2 diabetes in the United States. JAMA 2017;317(9):912–24. (eTable5) (MICHA et al., 2017) |
|  | Any other mortality (excluding CHD and stroke) | Individual level meta-analysis of 48 prospective cohort studies | Adjusted for age, sex, race or ethnicity, deprivation, smoking, diabetes, inactivity, alcohol, obesity | Stringhini S, Carmeli C, Jokela M, *et al*. Socioeconomic status and the 25 × 25 risk factors as determinants of premature mortality: a multicohort study and meta-analysis of 1·7 million men and women. The Lancet 2017;389(10075):1229–37. (Figure 4) (STRINGHINI et al., 2017) |
| Setting reference level of systolic blood pressure^1^ | Ideal systolic blood pressure below which no risk was considered | Evidence from evidence from randomised trials of antihypertensive drugs and the Intersalt study | There may be health benefits by lowering systolic blood pressure down to 110mmHg | Singh GM, Danaei G, Farzadfar F, *et al*. The age-specific quantitative effects of metabolic risk factors on cardiovascular diseases and diabetes: a pooled analysis. PLOS ONE 2013;8(7):e65174. (SINGH et al., 2013) |
| Disease costs | Hospitalisation costs for CHD and stroke | Based on the Medical Expenditure Panel Survey (MEPS) | Stratified by age and sex | Ministry of Health, National Hospital Information System (SIH/SUS – *Sistema de Informações Hospitalares*) - Underlying cause of hospitalization based on the ICD-10 codes. 2018 (MINISTÉRIO DA SAÚDE, 2017b) |
|  | Informal care costs for CHD |  | Costs were extrapolated for US settings | Leal J, Luengo-Fernández R, Gray A, Petersen S, Rayner M. Economic burden of cardiovascular diseases in the enlarged European Union. Eur Heart J 2006;27(13):1610–9. (Table 5) |

Table B Key modeling assumptions and limitations.

Population module

We assumed no migration after the age of 30

We assumed POF and PNS to be representative of the Brazilian population

Disease module

We assumed log-linear exposure – response relationship for SBP with 5-year mean lag time

We only modeled first ever event of CHD and stroke because we focus on primary prevention

For CHD and stroke initial incidence rates (year 2018), we used modeled estimates derived from mortality and PNS 2013 prevalence data

We assumed the non-attributable to SBP incidence rate trends for CHD and stroke, to be 50% of the forecasted mortality rates trends

We assumed that the risk ratios of SBP on CHD and stroke incidence and mortality are equal and SBP is not modifying CHD and stroke survival

We assumed that changes in sodium consumption have an immediate effect on SBP and changes in SBP have a median 5-year time lag to impact the risk of CVD

Policy module

We assumed that the recently observed trends in sodium consumption, SBP, and disease specific mortality would continue in the future (baseline scenario)

We assumed that the Brazilian population diet has and will have similar food composition since 2008-09

We assumed that individuals would not change sodium consumption behavior because of the policy

Table C. Food categories in the Brazilian voluntary agreements, the equivalent food categories in the POF 2008-2009 survey and their mean sodium content at the baseline of negotiations and in 2017 (mg/100g).

| Food description voluntary agreements | POF 2008-2009 Food category | Mean sodium content (mg/100g) | |
| --- | --- | --- | --- |
|  |  | Baseline | 2017 |
| Instant pasta (noodles) | Massas instantâneas | 1960.0 | 1598.6 |
| Sliced bread | Pão de forma | 426.5 | 365.0 |
| Buns | N/A | 436.1 | 374.4 |
| Cakes without filling | Bolo sem recheio | 335.7 | 241.1 |
| Cakes with filling | N/A | 249.9 | 185.8 |
| Creamy cake mixes^a^ | Mistura para bolo | 270.7 | 229.6 |
| Aerated cake mixes^a^ | Mistura para bolo | 372.3 | 291.6 |
| Potato chips | Batata frita, batata palha | 547.6 | 475.4 |
| Extruded corn snacks | Snacks | 831.9 | 827.4 |
| Filled cookies | Biscoito recheado | 259.5 | 235.5 |
| Salted crackers | Biscoito salgado | 695.8 | 590.9 |
| Sweet biscuits | Biscoitos doces | 359.5 | 293.9 |
| Mayonnaise | Maionese | 1063.3 | 852.7 |
| Dairy/cheese spread | Requeijão | 659.5 | 434.5 |
| Margarines | Margarina | 739.9 | 544.3 |
| Mozzarella cheese | Queijo muçarela | 600.2 | 517.2 |
| Rice condiments | Demais temperos | 31,425.1 | 31,260.0 |
| Bouillon cubes or powders^b^ | Caldo cubo, pó | 1,035.9 | 952.1 |
| Paste condiments | Tempero em pasta | 33,494.5 | 31,845.7 |
| Breakfast cereals | Cereais matinais | 428.9 | 359.2 |
| Soups | Sopa | 334.2 | 295.1 |
| Breaded meat | Empanados | 684.1 | 588.7 |
| Hotdog | Salsicha | 1,136.7 | 1,082.9 |
| Bologne | Mortadela | 1,328.0 | 1,435.5 |
| Sausage | Linguiça cozida | 1,323.0 | 1,210.8 |
| Hamburger | Hambúrguer | 816.6 | 630.2 |
| Fresh sausage | Linguiça frescal | 1,091.2 | 1,001.2 |
| Ham | Presuntaria | 1,203.9 | 1,186.6 |
| Sausage (stored at room temperature) | Linguiça cozida | 1,537.9 | 1,323.7 |
| French bread | Pão francês | 320.0 | 289.0 |

^a^ = as consumed

^b^= prepared according to label instructions

Table D. Sodium intake by age and sex groups and by dietary sodium sources at baseline (POF 2008-2009 survey – Personal Food Consumption Module, adjusted by the total sodium intake from the Food Acquisition Module).

|  | Added salt | | | | Other sodium sources | | | |
| --- | --- | --- | --- | --- | --- | --- | --- | --- |
|  | Mean | SE | CI – 95% |  | Mean | SE | CI – 95% |  |
| Men |  |  |  |  |  |  |  |  |
| 30-34y | 7,674.42 | 168.31 | 7,344.52 | 8,004.33 | 6,312.18 | 183.22 | 5,953.07 | 6,671.30 |
| 35-39y | 7,727.56 | 192.82 | 7,349.62 | 8,105.49 | 6,450.00 | 197.95 | 6,062.02 | 6,837.98 |
| 40-44y | 7,712.25 | 175.23 | 7,368.80 | 8,055.70 | 5,966.45 | 166.95 | 5,639.21 | 6,293.69 |
| 45-49y | 7,312.35 | 163.53 | 6,991.83 | 7,632.87 | 6,257.26 | 265.73 | 5,736.43 | 6,778.09 |
| 50-54y | 7,671.73 | 199.76 | 7,280.19 | 8,063.27 | 5,575.10 | 192.52 | 5,197.75 | 5,952.45 |
| 55-59y | 7,095.57 | 191.30 | 6,720.62 | 7,470.51 | 5,814.69 | 239.13 | 5,346.00 | 6,283.39 |
| 60-64y | 6,965.55 | 203.01 | 6,567.64 | 7,363.47 | 5,738.10 | 271.43 | 5,206.09 | 6,270.11 |
| 65-69y | 6,586.52 | 271.70 | 6,024.38 | 7,148.66 | 6,104.86 | 251.83 | 5,583.82 | 6,625.89 |
| Women |  |  |  |  |  |  |  |  |
| 30-34y | 5,378.36 | 137.51 | 5,108.84 | 5,647.88 | 5,344.74 | 146.89 | 5,056.84 | 5,632.64 |
| 35-39y | 5,402.00 | 109.97 | 5,186.45 | 5,617.54 | 5,324.91 | 139.50 | 5,051.48 | 5,598.34 |
| 40-44y | 5,401.88 | 103.12 | 5,199.76 | 5,604.00 | 5,139.15 | 132.50 | 4,879.45 | 5,398.85 |
| 45-49y | 5,141.73 | 113.49 | 4,919.28 | 5,364.18 | 5,175.08 | 134.35 | 4,911.74 | 5,438.42 |
| 50-54y | 5,089.95 | 143.58 | 4,808.53 | 5,371.37 | 5,309.76 | 195.51 | 4,926.56 | 5,692.96 |
| 55-59y | 5,213.10 | 134.97 | 4,948.56 | 5,477.64 | 5,080.89 | 156.52 | 4,774.10 | 5,387.68 |
| 60-64y | 5,136.61 | 150.62 | 4,841.38 | 5,431.83 | 4,878.31 | 168.20 | 4,548.63 | 5,207.99 |
| 65-69y | 4,780.91 | 165.98 | 4,455.57 | 5,106.24 | 5,403.88 | 187.61 | 5,036.15 | 5,771.60 |

Table E. Estimated sodium intake by age and sex groups and by dietary sodium sources considering food reformulation caused by the Brazilian voluntary target scenarios (2017).

|  | Added salt | | | | Other sodium sources | | | |
| --- | --- | --- | --- | --- | --- | --- | --- | --- |
|  | Mean | SE | CI – 95% |  | Mean | SE | CI – 95% |  |
| Men |  |  |  |  |  |  |  |  |
| 30-34y | 6,312.18 | 183.22 | 5,953.07 | 6,671.30 | 6,062.69 | 175.98 | 5,717.77 | 6,407.61 |
| 35-39y | 6,450.00 | 197.95 | 6,062.02 | 6,837.98 | 6,200.51 | 190.29 | 5,827.53 | 6,573.48 |
| 40-44y | 5,966.45 | 166.95 | 5,639.21 | 6,293.69 | 5,716.96 | 159.97 | 5,403.40 | 6,030.51 |
| 45-49y | 6,257.26 | 265.73 | 5,736.43 | 6,778.09 | 6,007.77 | 255.13 | 5,507.70 | 6,507.83 |
| 50-54y | 5,575.10 | 192.52 | 5,197.75 | 5,952.45 | 5,325.60 | 183.91 | 4,965.14 | 5,686.07 |
| 55-59y | 5,814.69 | 239.13 | 5,346.00 | 6,283.39 | 5,565.20 | 228.87 | 5,116.61 | 6,013.79 |
| 60-64y | 5,738.10 | 271.43 | 5,206.09 | 6,270.11 | 5,488.60 | 259.63 | 4,979.72 | 5,997.48 |
| 65-69y | 6,104.86 | 251.83 | 5,583.82 | 6,625.89 | 5,855.36 | 241.54 | 5,355.62 | 6,355.10 |
| Women |  |  |  |  |  |  |  |  |
| 30-34y | 5,344.74 | 146.89 | 5,056.84 | 5,632.64 | 5,095.24 | 140.03 | 4,820.78 | 5,369.71 |
| 35-39y | 5,324.91 | 139.50 | 5,051.48 | 5,598.34 | 5,075.42 | 132.97 | 4,814.80 | 5,336.04 |
| 40-44y | 5,139.15 | 132.50 | 4,879.45 | 5,398.85 | 4,889.66 | 126.06 | 4,642.57 | 5,136.74 |
| 45-49y | 5,175.08 | 134.35 | 4,911.74 | 5,438.42 | 4,925.59 | 127.88 | 4,674.95 | 5,176.23 |
| 50-54y | 5,309.76 | 195.51 | 4,926.56 | 5,692.96 | 5,060.27 | 186.32 | 4,695.07 | 5,425.46 |
| 55-59y | 5,080.89 | 156.52 | 4,774.10 | 5,387.68 | 4,831.40 | 148.84 | 4,539.67 | 5,123.12 |
| 60-64y | 4,878.31 | 168.20 | 4,548.63 | 5,207.99 | 4,628.82 | 159.60 | 4,315.99 | 4,941.64 |
| 65-69y | 5,403.88 | 187.61 | 5,036.15 | 5,771.60 | 5,154.39 | 178.95 | 4,803.64 | 5,505.13 |

#### Additional result from main analysis

Table F Health related model estimates over the 20-year simulation period from 2013 to 2032, for Brazilian adults age 30 to 84 years by sex. Values are the median estimate (95% UI). Negative costs represent savings.

| Age group | Sex | All causes | CHD | Stroke | Non-CVD causes |
| --- | --- | --- | --- | --- | --- |
| Deaths prevented or postponed (DPP) |  |  |  |  |  |
| 30-49y | Men | 918 (229-2293) | 0 (0-688) | 0 (0-460) | 688 (0-1836) |
|  | Women | 230 (0-1146) | 0 (0-230) | 0 (0-459) | 229 (0-689) |
| 50-69y | Men | 4,586 (2069-7571) | 459 (0-1378) | 460 (0-1605) | 3,440 (1376-6193) |
|  | Women | 2,525 (919-4815) | 229 (0-918) | 458 (0-1147) | 1,836 (460-3679) |
| >=70y | Men | 3,213 (1375-5736) | 229 (-230-918) | 230 (-229-1148) | 2,752 (917-4819) |
|  | Women | 3,212 (1377-5515) | 229 (-229-918) | 230 (-229-1147) | 2,525 (918-4819) |
| All ages | Men | 8,721 (5047-13765) | 917 (0-2293) | 920 (229-2522) | 6,879 (3440-11241) |
|  | Women | 6,194 (3212-9870) | 459 (0-1606) | 916 (0-2065) | 4,816 (2292-8042) |
| Cases prevented or postponed (CPP) |  |  |  |  |  |
| 30-49y | Men |  | 12,390 (1836-36032) | 4,816 (230-14221) |  |
|  | Women |  | 2,521 (0-11467) | 3,441 (0-11701) |  |
| 50-69y | Men |  | 42,212 (13308-92696) | 27,758 (8945-58737) |  |
|  | Women |  | 16,519 (3443-39688) | 22,940 (6885-48642) |  |
| >=70y | Men |  | 12,388 (1831-28447) | 12,387 (2294-27996) |  |
|  | Women |  | 12,157 (2753-27072) | 12,847 (3442-28678) |  |
| All ages | Men |  | 68,134 (24321-149346) | 45,198 (15142-95902) |  |
|  | Women |  | 31,890 (9176-71584) | 39,463 (13995-85337) |  |
